# Extending the Depth of LED-based Photoacoustic Imaging for Carotid and Breast Applications

**DOI:** 10.1101/2025.11.13.25339983

**Authors:** Anjali Thomas, Mithun Kuniyil Ajith Singh, Naoto Sato, Kalloor Joseph Francis

**Author notes:** Corresponding author: Kalloor Joseph Francis. Anjali Thomas and Mithun Kuniyil Ajith Singh contributed equally to this work.

## Abstract

Light-emitting diode (LED)-based photoacoustic (PA) imaging offers a compact, safe, and cost-effective alternative to laser systems. However, low LED power leads to low optical fluence, which in turn limits penetration depth. We report a systematic optimization of LED-PA performance by jointly tuning ultrasound (US) probe frequency and LED pulse width, validated in both phantom and in vivo studies. Using a commercially available LED-PA/US platform, we compared a custom 5 MHz transducer with commercial 7 and 10 MHz probes under LED pulses of 30 to 100 ns. The 5 MHz probe with a 100 ns pulse achieved the best trade-off between depth sensitivity and resolution, enabling detection of targets up to 18 mm. In vivo experiments demonstrated, for the first time, clear visualization of the carotid artery and deep-seated breast vessels using LED-based PA imaging. These findings show that careful optimization of probe frequency and pulse width can substantially extend the depth performance of LED-PA, advancing its potential for vascular and oncologic applications.

## I. Introduction

Light-emitting diode (LED)-based photoacoustic (PA) imaging has emerged as a compact, cost-effective, and intrinsically safe alternative to conventional laser-based systems [1], [2]. Unlike bulky and expensive laser platforms, LEDs are portable, energy-efficient, and capable of high pulse repetition rates, making them well-suited for clinical translation and point-of-care use. These advantages are particularly relevant for applications such as breast cancer imaging and carotid plaque assessment, where real-time, bedside imaging could provide significant diagnostic value [3], [4]. However, a major challenge of LED-based PA imaging is its inherently low optical pulse energy, which limits light penetration into tissue and consequently constrains imaging depth. Current in-vivo studies with commercially available LED-PA systems have typically achieved penetration depths of only about 10 mm, restricting their use to superficial vasculature [5]. For LED-PA to address deeper clinical targets, such as tumor-associated breast vasculature or atherosclerotic carotid plaques, it is essential to optimize both the optical excitation and the acoustic detection. In particular, maximizing LED pulse energy and matching its spectrum and temporal profile to the ultrasound (US) transducer bandwidth are critical to strengthen PA signal and extend imaging depth [6], [7], [8]. Lengthening the LED pulse can increase emitted energy, provided thermal and stress confinement are preserved, and the US detector must retain sensitivity across the resulting PA frequency band. Here, we systematically investigate these trade-offs for deep tissue imaging.

We introduce a custom-developed 5 MHz transducer designed to capture the low-frequency spectrum critical for deep-tissue photoacoustic imaging. This design enables the use of longer LED pulses (100 ns), which substantially increase optical output while maintaining spectral compatibility with the transducer’s frequency response. We systematically evaluated this approach in ex vivo tissue phantoms using 5 MHz, 7 MHz, and 10 MHz ultrasound probes with 30 ns to 100 ns LED pulses. With the optimized 5 MHz and 100 ns configuration, we demonstrate for the first time in vivo LED-PA imaging of the human carotid artery and deep-seated breast vessels at depths up to 18 mm. This advancement establishes a clear pathway toward clinically relevant applications in carotid plaque characterization and breast cancer imaging. In the carotid, penetration beyond 1 cm is needed to resolve the vessel wall for plaque assessment and stroke risk evaluation. In the breast, tumor-associated and feeding vessels are commonly located 10 to 30 mm from the skin and can extend deeper depending on breast composition and lesion site [9], [10]. This depth range is also relevant for carotid artery imaging [6], [7]. Hence, meeting these depth ranges with an LED-based system represents a key step toward point-of-care vascular and oncologic imaging, even in resource-limited settings.

## II. Methods

We used a commercially available LED-based PA/US imaging system (AcousticX, CYBERDYNE Inc., Japan) equipped with dual 850 nm LED arrays and interchangeable linear probes. The system allows adjustment of LED pulse widths between 30 and 100 ns, corresponding to measured optical energies between 118 and 498 *µ*J. Increasing the pulse duration substantially enhances optical output, which is critical for deep-tissue imaging. Under stress confinement, the axial resolution is approximately the product of pulse duration and the speed of sound in tissue (∼1500 m/s). For a 100 ns pulse, this corresponds to ∼ 150 *µ*m. A target of this dimension predominantly generates photoacoustic signals with a center frequency given by

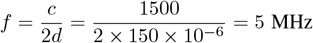

where *c* is speed of sound and *d* is the diameter of the target. We developed a custom 5 MHz linear array transducer to match the expected spectral content of signals generated under 100 ns excitation. Even though the achievable spatial resolution is limited by the probe reception bandwidth, this design allows us to harness the higher optical energy of longer LED pulses without spectral mismatch, achieving an optimal trade-off between penetration depth and resolution. To validate this configuration, we compared three linear ultrasound transducers (Table I) under different LED pulse durations to assess frequency matching and depth-dependent performance.

**TABLE I:** Transducer specifications.

| Probe | No of elements | Pitch (mm) | Element size (mm) | Focus depth (mm) | Bandwidth (-6dB) (MHz) | Fractional bandwidth (%) |
| --- | --- | --- | --- | --- | --- | --- |
| 5 MHz | 96 | 0.6 | 10 | 30 | 3 - 7 | 83.5 |
| 7 MHz | 128 | 0.315 | 4 | 20 | 4 - 10 | 77.5 |
| 10 MHz | 128 | 0.3 | 3.5 | 15 | 5 - 12 | 85 |

To characterize the optimal configuration, we performed ex vivo measurements using chicken breast tissue with embedded silicone tube (inner diameter 4 mm) filled with titanium ink and embedded at a depth of 29 mm. The tube was imaged in cross-section with all three transducers coupled to the 850 nm LED arrays, using pulse widths of 30, 60, and 100 ns (Fig. 1a). Ultrasound gel was applied to ensure acoustic coupling between the probe and the tissue surface. Data acquisition was performed at a pulse repetition frequency of 4 kHz. A total of 384 frames were averaged in real time, resulting in an effective frame rate of 6.25 Hz. The tube position was first identified using B-mode ultrasound and maintained at the same location for all measurements. For each probe - pulse configuration, both PA and US RF data were acquired and stored for offline processing. The PA signal from the tube, recorded with the transducer element directly above it, was used to analyze the frequency response.

**Fig. 1.**
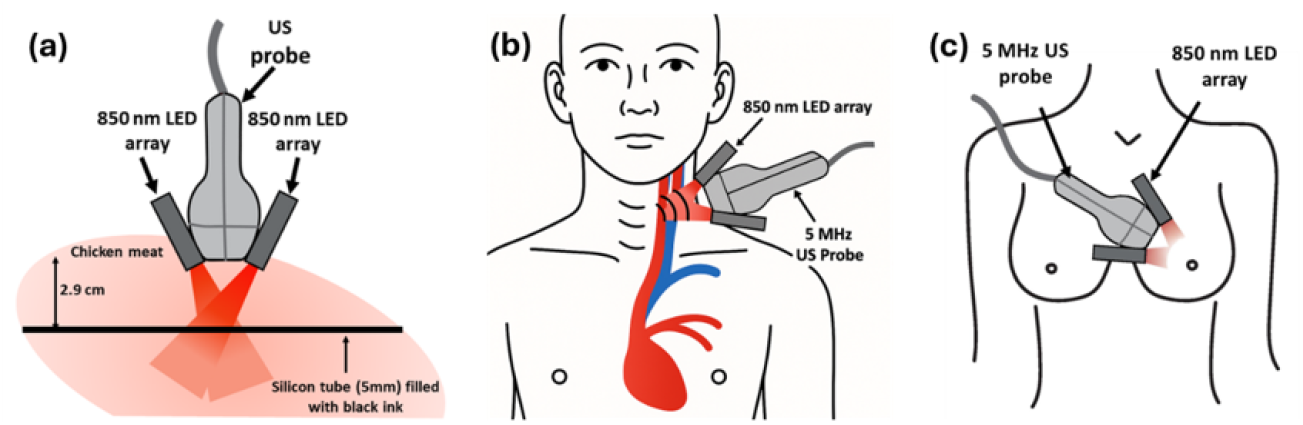
Experimental setups for LED-based photoacoustic imaging. (a) Ex vivo phantom: silicone tube (4 mm ID) filled with titanium ink and embedded at 29 mm depth in chicken tissue. (b) In vivo carotid imaging with the probe positioned on the neck over the carotid artery and jugular vein. (c) In vivo breast imaging with the probe and LED arrays placed on the breast surface using a gel pad for acoustic and optical coupling.

The optimized configuration for deep vasculature imaging was evaluated in vivo. For carotid artery and jugular vein imaging, adult female volunteer (Fitzpatrick type II) was examined in the sitting position (Fig. 1b). Line-by-line ultrasound acquisition was combined with PA imaging to improve image quality. Breast imaging was performed on a second volunteer (adult female, Fitzpatrick type III) using the same setup (Fig. 1c), with the probe placed on the breast surface over a gel pad to ensure both acoustic and optical coupling. Written informed consent was obtained from volunteers before imaging in accordance with the study protocol MEC-2014-611.

## III. Results and discussion

Fig. 2(a–i) show PA/US overlays from the ex vivo phantom. Grayscale encodes US anatomy and the hot colormap encodes PA amplitude, which helps localize absorbers in tissue. The titanium-ink tube at 29 mm depth was visualized with all three probes, but sensitivity depended strongly on probe frequency and LED pulse width. The 5 MHz probe consistently produced the highest PA signal, with a marked increase as the pulse width was extended to 100 ns, in line with the measured rise in optical output from 118 to 498 *µ*J between 30 and 100 ns. PA signals from the center element in Fig. 2j show that the 5 MHz probe yields the highest target signal, which increases with pulse duration. In contrast, the 7 and 10 MHz transducers record PA amplitudes that are 8–10 times lower. These results indicate that the 5 MHz transducer benefits most from longer pulse widths within the frequency range examined.

**Fig. 2.**
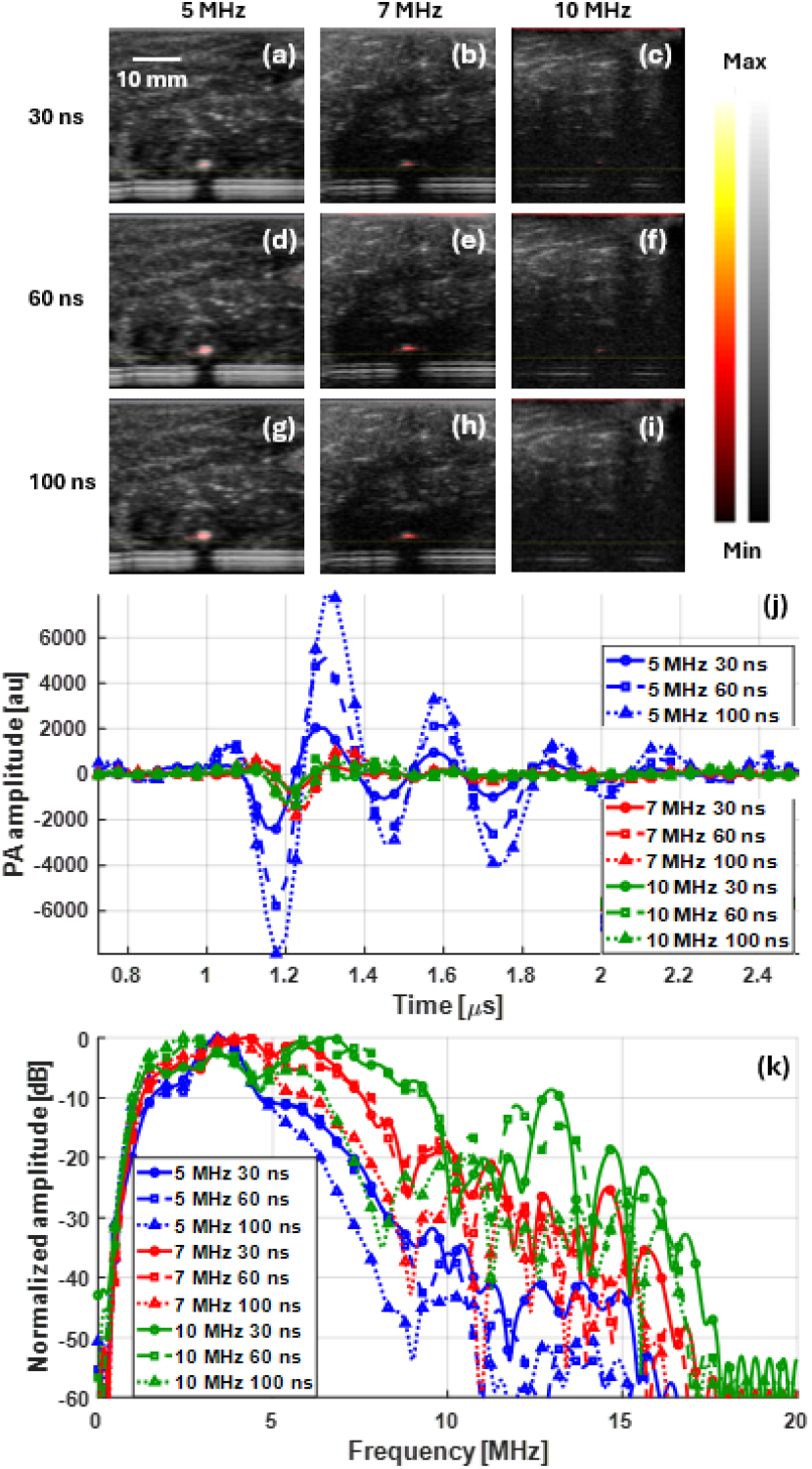
Ex vivo phantom imaging results. (a–i) PA/US cross-sectional images of a silicone tube (4 mm ID, 2.9 cm depth) filled with titanium ink acquired with 5, 7, and 10 MHz probes at 30, 60, and 100 ns pulse widths. (j) Quantified peak PA amplitude as a function of probe frequency and pulse width, showing the strongest response with the 5-MHz probe at 100 ns. (k) Normalized frequency response of the detected photoacoustic signal. reception, enabling robust signal detection from depths beyond 20 mm.

The frequency response in Fig. 2k shows that the dominant spectral content from the 29 mm deeper target lies in the 3-6 MHz band, which aligns with the 5 MHz transducer. By contrast, the 7 and 10 MHz probes sample near the edges of their passbands at this depth, which increases attenuation and reduces effective sensitivity. This frequency matching explains the 5 MHz advantage at 100 ns, where longer pulses deliver higher optical energy without incurring a spectral mismatch. Moreover, the additional low frequency components generated at longer pulse widths remain within the sensitive band of the 5-MHz probe, but fall outside the optimal response of the higher frequency probes. As a result, the 5-MHz configuration uniquely combines improved fluence with favorable acoustic. reception, enabling robust signal detection from depths beyond 20 mm.

At 100 ns, resolution measurements with the 5 MHz probe yielded 470 *µ*m axial and 700 *µ*m lateral FWHM, which is coarser than the sub-200 *µ*m expected with 10 MHz at shallow depth, yet sufficient for millimeter-scale vascular targets such as dilated tumor-associated vessels in the breast and the jugular and carotid vessels. This represents a practical trade-off, fine enough to capture vessel morphology while delivering the sensitivity and penetration required for clinically relevant depths in breast and neck imaging.

Fig. 3(a–d) shows in vivo imaging results of the carotid region with the optimized 5 MHz probe and 100 ns LED pulses. A real-time video corresponding to Fig. 3(c) and (d) are provided in the supplementary material. Despite overlying muscle, optically absorbing skin, and subcutaneous fat, the carotid artery is clearly visualized. PA highlights intraluminal blood, and US provides complementary anatomy for localization. The upper carotid wall is detected at a depth of about 9 mm, which matches the expected anatomy. Frame averaging stabilized the overlay and improved SNR while preserving alignment. The internal jugular vein is visible in the same field of view and appears partially compressed in the seated posture. These results indicate that LED-PA can add vascular contrast to US in clinically relevant neck arteries, supporting future work on plaque characterization and vessel-wall assessment at depth.

**Fig. 3.**
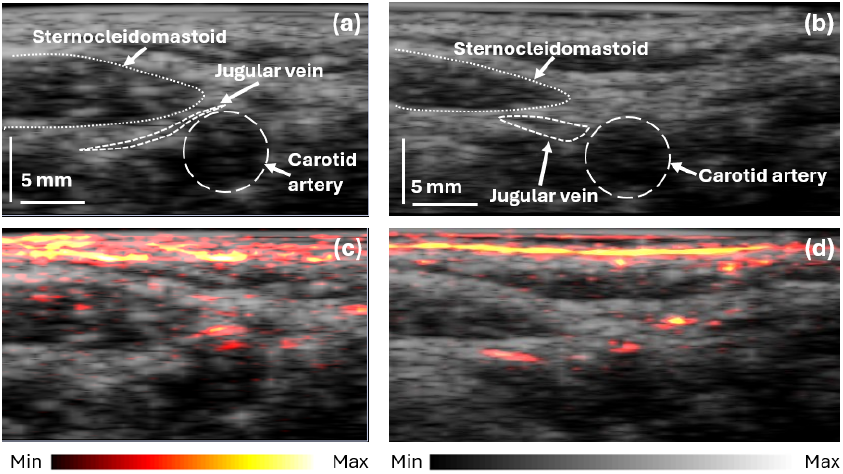
In vivo PA/US imaging of cervical vasculature using the optimized 5-MHz probe with 100-ns LED pulses. (a, b) US-only image, and (c, d) PA/US overlay showing carotid artery and jugular vein. Corresponding real-time videos are provided in the supplementary material. Colormap indicates PA amplitude (hot), overlaid on grayscale ultrasound anatomy.

Fig. 4(a–d) show in vivo breast imaging with the optimized 5 MHz transducer and 100 ns pulse width. Figures 4a and 4b are ultrasound images from two locations. Figures 4c and 4d are PA/US overlays that reveal blood vessels. Both superficial vessels near the skin and deeper vessels down to 18 mm are visible. The PA signal highlights vascular channels that are less distinct on ultrasound alone, providing complementary functional contrast. These depths are clinically relevant, since tumor-associated angiogenesis often involves dilated or tortuous vasculature that can extend from superficial layers into deeper feeding vessels. Detecting vasculature approaching 20 mm with an LED-based system is a notable advance and underscores the potential of this modality for characterizing tumor microenvironments and guiding breast cancer diagnostics in resource-constrained settings.

**Fig. 4.**
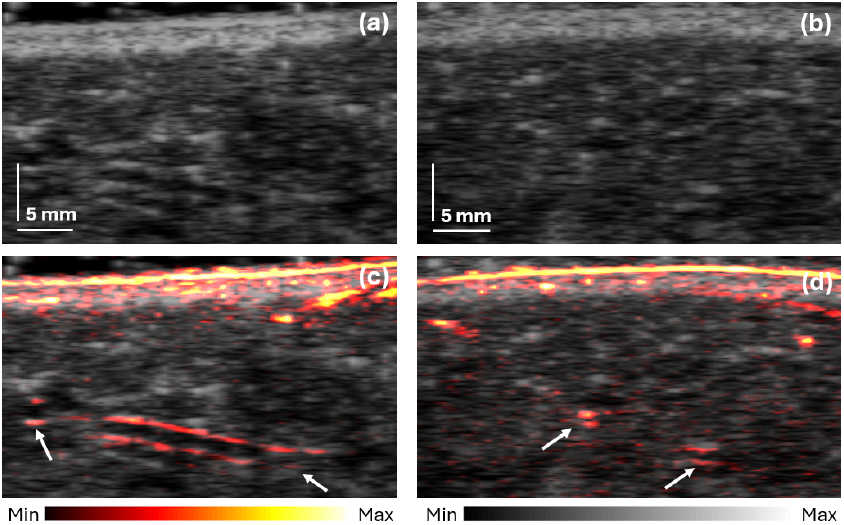
In vivo PA/US imaging of breast using the optimized 5-MHz probe with 100-ns LED pulses. (a, b) US-only image, and (c, d) PA/US overlay demonstrating visualization of superficial and deep vessels. Colormap indicates PA amplitude in hot, overlaid on grayscale ultrasound anatomy.

Beyond the technical optimization demonstrated here, the broader significance of LED-based PA imaging lies in its potential for accessible clinical adoption. Unlike laser systems, LED sources are compact, affordable, and intrinsically eye-safe, enabling integration into portable platforms. Combined with co-registered US, LED-PAI provides both anatomical and functional vascular information in a single handheld device. These attributes make it particularly attractive for carotid and breast applications, where rapid, bedside assessment of vascular structures could support early diagnosis and monitoring. By reducing cost and complexity while maintaining clinical relevance, LED-PAI represents a practical pathway toward widespread, point-of-care use in settings where conventional PA systems would be impractical.

While this study demonstrates clear progress in extending the imaging depth of LED-based photoacoustics, the achieved penetration remains at the threshold of what is required for consistent clinical use. Validation in larger volunteer groups will be essential to establish reliability across patient populations. Our current work demonstrates feasibility, and ongoing efforts are directed toward improving system compatibility and data acquisition sensitivity to further enhance depth performance.

Future developments should also prioritize multispectral capability, or at least dual-wavelength operation. In the carotid, this would allow assessment of oxygenation and lipid content relevant to plaque vulnerability, while in the breast, it could provide functional insight into tumor angiogenesis and hypoxia. Adding multispectral capability would extend LED-PA from anatomical vascular imaging toward functional and molecular characterization, broadening its clinical utility.

## IV. Conclusion

In this study, we optimized an LED-based PA imaging system for clinically relevant depths by pairing a sensitive 5 MHz transducer with 100 ns LED pulses. This configuration delivered approximately 0.5 mJ per pulse per array, to our knowledge the highest energy reported for LED-driven PA imaging, and aligned the generated PA spectrum with the transducer passband. Ex vivo experiments in chicken tissue visualized targets at 29 mm. In vivo, we achieved real-time PA/US imaging of the carotid artery at about 9 mm and breast vasculature up to 18 mm. These results demonstrate that selection of probe frequency and pulse duration extends the depth reach and improves sensitivity of LED-PA. The approach is applicable to vascular assessments in peripheral artery and breast imaging, with potential to aid plaque characterization and interrogation of tumor-associated vasculature. By leveraging compact, low-cost, and intrinsically safe LED sources integrated with ultrasound, this modality offers a practical route to point-of-care deployment and can broaden access to vascular and oncologic imaging outside specialized centers.

## Supporting information

Video file for Fig_3c

Video file for Fig_3d

## Data Availability

All data produced in the present study are available upon reasonable request to the authors.

